# CHARACTERIZATION OF ACROMEGALY ACCORDING TO TUMOR SIZE AT DIAGNOSIS

**DOI:** 10.1101/2024.03.19.24304552

**Authors:** Leticia Marinho Del Corso, Vicente Florentino Castaldo Andrade, Solena Ziemer Kusma Fidalski, Cesar Luiz Boguszewski

**Affiliations:** SEMPR (Endocrine Division), Department of Internal Medicine, Federal University of Parana, Curitiba, Brazil; Department of Public Health, Federal University of Parana, Curitiba, Brazil

**Keywords:** acromegaly, pituitary adenoma, growth hormone-secreting pituitary adenoma, giant tumor

## Abstract

**Purpose:** To evaluate the clinical, laboratory, radiological, therapeutic, and prognostic characteristics of patients with acromegaly according to the size of the growth hormone (GH)-secreting pituitary adenoma at diagnosis.

**Patients and Methods:** Observational, retrospective, single-center study of patients with acromegaly followed at a tertiary center. Data from medical records were evaluated regarding age, symptoms, presence of arterial hypertension, type 2 diabetes mellitus, hypopituitarism, size of the initial lesion, invasiveness (cavernous sinus invasion), T2-weighted magnetic resonance imaging signal intensity, GH and insulin-like growth factor type 1 (IGF-1) levels, treatment performed [surgery, use of somatostatin receptor ligands (SRL), pegvisomant, cabergoline and bromocriptine and radiotherapy] and response to surgical or adjuvant treatment (normal levels of GH and/or IGF-1 after each treatment instituted).Patients were divided into groups according to the size of the adenoma at diagnosis (group I = ≤ 10 mm, II = 10-19 mm, III = 20-29 mm, IV = 30-39 mm and V = ≥ 40 mm), and comparisons were made between the 5 groups and two-by-two comparisons.

**Results:** 117 patients were studied (59 women, age at diagnosis 43 ± 13 years). Group I consisted of 11 patients (9%), group II of 54 (46%), group III of 34 (29%), group IV of 10 (9%) and group V of 8 patients (7%). The prevalence of hypertension, diabetes mellitus and hypopituitarism were 49%, 25% and 28%, respectively. Hypopituitarism, invasiveness, and the use of SRL had their prevalence increased according to the size of the adenoma, as well as GH levels. Age, on the other hand, showed a negative correlation with tumor size, and group I was older when compared to the group with macroadenoma. The ROC curves showed that in relation to the size of the adenoma at diagnosis, most of the outcomes evaluated (hypopituitarism, invasiveness, radiotherapy, use of SRL, use of medications other than SRL, disease control after surgery) occurred with a tumor diameter of around 20 mm.

**Conclusion:** Our study demonstrated that microadenomas and macroadenomas < 20 mm are associated with lower morbidity and better therapeutic response in acromegaly. From a tumor diameter of 20 mm, there was no significant difference in the clinical, therapeutic and prognostic behavior of GH-secreting pituitary adenomas.

**Trial Registration number (Plataforma Brasil):** CAAE 30066220.2.0000.0096 (April 02, 2020)

## INTRODUCTION

Acromegaly is a chronic and insidious disease characterized by increased circulating levels of growth hormone (GH) and insulin-like growth factor-I (IGF-I), in most cases caused by a GH-secreting pituitary adenoma [1]. At diagnosis, approximately 75% of patients with acromegaly have a macroadenoma, which is defined by a tumor measuring 10 mm (1 cm) or more in its larger diameter. It is estimated that less than 5% of patients present larger adenomas called “giant tumors”, which have been defined as those having more than 40 mm (4 cm) in their largest diameter [2 - 4]. Giant macroadenomas are usually considered to be more invasive, cause greater morbidity, present less chance to be completely removed by surgery, and have worse therapeutic responses and prognosis [3 - 5]. However, the widely adopted cutoff point of 4 cm was taken arbitrarily, as there have been no studies evaluating whether this cutoff is the best measurement to show differences in relation to the clinical presentation, behaviour and prognosis of these tumors compared to smaller ones [6 - 8].

The present study aimed to evaluate clinical, laboratory, imaging, therapeutic management, and prognostic characteristics of patients with acromegaly according to the size of the GH-secreting pituitary adenoma at the time of the diagnosis of the disease, and to investigate from which cutoff point these outcomes do not differ significantly.

## PATIENTS AND METHODS

### Study design

This is an observational, retrospective, single-center study of patients with the diagnosis of acromegaly (ICD-10 E22.0), who were or are still being followed up at our institution (HC-UFRP/SEMPR), which is a reference center for pituitary diseases in South of Brazil. The study was approved by the Human Research Ethics Committee of our institution. Patients were included if they were 18 years or older at the time of the analysis, and if they had the description of the tumor size at diagnosis. Patients were excluded from the analysis when data regarding the size of the tumor at diagnosis was not available and if acromegaly was not caused by a pituitary adenoma.

We have evaluated and collected data from medical records, including patient’s age at diagnosis, time elapsed between the first signs and/or symptoms until the diagnosis, presence of arterial hypertension (AH), type 2 diabetes (DM) and hypopituitarism, GH and IGF-1 levels [also in percentual above the upper limit of normal (ULN)] at diagnosis, characteristics in the magnetic resonance imaging (MRI) such as size of the GH-secreting pituitary adenoma, invasiveness (according to Knosp classification when possible), and T2-weighted MRI signal intensity, therapeutic approach [surgery, radiotherapy, or medical therapy with somatostatin receptor ligands (SRL), cabergoline (CBG) and/or GH-receptor antagonist pegvisomant (PEGV)], and prognosis in relation if disease control was obtained with surgery, adjuvant therapies or was not controlled.

The study group was categorized according to the size of the GH-secreting pituitary adenoma at diagnosis: Group I, < 10mm (microadenomas); Group II, between 10-19 mm; Group III, between 20-29 mm; Group IV, between 30-39 mm; Group V, ≥ 40 mm (giant). The information found in the five groups was analyzed and compared and it was also performed an analysis comparing them two by two: adenomas < 10 mm vs ≥ 10 mm; Group II vs Group III, IV and V; Group III vs Group IV and V; Group IV vs Group V.

### Statistical analysis

The data collected from the diaries were transferred to IBM SPSS Statistics 21 version software spreadsheet. The normality of the variables was assessed using the Shapiro-Wilk test. Results are described as means plus minus standard deviations (SD), medians, minimum and maximum values (quantitative variables) or by frequencies and percentages (categorical variables). To compare variables with normal distribution, the Student’s t and ANOVA tests were applied, while the Mann-Whitney and Kruskal-Wallis tests were used for variables with asymmetric distribution. The comparisons between groups in relation to categorical variables were performed using the Chi-square test and Fisher’s exact test. For correlations between categorical variables, Spearman’s rho was used. A ROC Curve analysis was performed with the aim of evaluating the sensitivity and specificity of tumor size in predicting the various clinical outcomes studied. Two-sided p values < 0.05 were used to indicate significant statistical differences.

## RESULTS

We have evaluated 166 medical records, of which 49 were excluded. The characteristics of the study group are summarized in **Table 1**. The final study group consisted of 117 individuals: 59 women and 58 men, with a mean age at diagnosis of 43 ± 13 years. In the study group the median time from symptoms to diagnosis was 4 years (range 0.2-35 years). The prevalence of hypertension was 49% and of type 2 diabetes mellitus was 25%, while the overall prevalence of hypopituitarism was 28%. In relation to the the adenoma size at diagnosis, 11 patients (9%) had tumors smaller than 10 mm, 54 (46%) between 10-19 mm, 34 (29%) between 20-29 mm, 10 (9%) between 30-39 mm and 8 (7%) equal or greater than 40 mm. Hypointense, isointense and hyperintense adenomas on T2-weighted MRI was observed in 44%, 40% and 16% of the cases, respectively. Only 9 patients underwent radiotherapy (8%). Medical therapy with somatostatin receptor ligand (SRLs) was used in 74 patients of the study group: 49 (66,2%) octreotide, 14 (19%) lanreotide, 9 (12%) octreotide or lanreotide at different times, 1 (1,4%) pasireotide and 1 (1,4%) paltusotine. SRLs were used as primary therapy in 22 (30%) and as adjuvant therapy in 48 (65%) of the patients. In 39 (34%) of patients other medications were needed: cabergoline in 28 (71,8%) patients, pegvisomant in 2 (5,1%), combined cabergoline and pegvisomant in 6 (15,4%) and bromocriptine in 3 (7,7%). The median GH was 13.4 ng/ml (range 0.52-135); median IGF-1 was 734 ng/ml (range 341-2.197) and median IGF-1/ULN was 2.73 times (range 1.25-6.14). Good prognosis with biochemical control of acromegaly was observed in 67% (70 patients out of a total of 105 with this data available). Of the patients with controlled disease, 29 (41%) had remission with surgery, 9 (13%) after primary drug treatment, 25 (36%) after combined treatment with surgery and medication and 7 (10%) after combined treatment with surgery, medication and radiotherapy. Poor prognosis due to lack of disease control was observed in 35 (33%) of the 105 patients evaluated. The rate of remission with surgery, that is, patients who had primary treatment with surgery and had the disease controlled, occurred in 29 of 84 patients (35%).

**Table 1.** Characterization of patients with acromegaly, divided into groups in relation to tumor size at diagnosis: < 10 mm (Groups I), 10-19 mm (Group II), 20-29 mm (Group III), 30-39 mm (Group IV) and ≥ 40 mm (Group V) size at diagnosis.

| Feature | Total<br>n=117 | Group I<br>n=11 | Group II<br>n=54 | Group III<br>n=34 | Group IV<br>n=10 | Group V<br>n=8 | p<br>Value |
| --- | --- | --- | --- | --- | --- | --- | --- |
| Gender, n (%) |  |  |  |  |  |  | NS |
| Men | 58/117<br>(50%) | 5/11<br>(45%) | 22/54<br>(41%) | 20/34<br>(59%) | 7/10<br>(70%) | 4/8 (50%) |  |
| Women | 59/117<br>(50%) | 6/11<br>(55%) | 32/54<br>(59%) | 14/34<br>(41%) | 3/10<br>(30%) | 4/8 (50%) |  |
| Age (years) | 43 ± 13 | 51 ± 10 | 43 ± 12 | 41 ± 14 | 43 ± 16 | 42 ± 11 | NS |
| Time of symptoms (yrs) | 4<br>(0.2-35) | 3<br>(1-15) | 4,5<br>(1-30) | 4<br>(0.5-35) | 3<br>(0.2-10) | 3<br>(1-19) | NS |
| Hypertension<br>n (%) | 57/117<br>(49%) | 6/11<br>(55%) | 27/54<br>(50%) | 17/34<br>(50%) | 3/10<br>(30%) | 4/8 (50%) | NS |
| Diabetes<br>n (%) | 29/117<br>(25%) | 3/11<br>(27%) | 11/54<br>(20%) | 12/34<br>(35%) | 2/10<br>(20%) | 1/8 (13%) | NS |
| Hypopituitarism<br>n (%) | 31/111<br>(28%) | 0/11<br>(0%) | 5/51<br>(10%) | 15/32<br>(47%) | 5/10<br>(50%) | 6/7 (86%) | 0.000 |
| Invasiveness<br>n (%) | 45/111<br>(41%) | 0/11<br>(0%) | 17/52<br>(33%) | 16/33<br>(48%) | 6/9<br>(67%) | 6/7 (86%) | 0.002 |
| T2 signal<br>intensity<br>n (%) |  |  |  |  |  |  | NS |
| Hypointense | 30/68<br>(44%) | 3/5<br>(60%) | 17/27<br>(63%) | 7/23<br>(30%) | 0/6<br>(0%) | 3/7 (43%) |  |
| Isointense | 27/68<br>(40%) | 2/5<br>(40%) | 8/27<br>(30%) | 10/23<br>(44%) | 5/6<br>(83%) | 2/7 (29%) |  |
| Hyperintense | 11/68<br>(16%) | 0/5<br>(0%) | 2/27<br>(7%) | 6/23<br>(26%) | 1/6<br>(17%) | 2/7 (29%) |  |
| Radiotherapy<br>n (%) | 9/117<br>(8%) | 0/11<br>(0%) | 1/54 (2%) | 5/34<br>(15%) | 1/10<br>(10%) | 2/8 (25%) | NS |
| SRL<br>n (%) | 74/117<br>(63%) | 3/11<br>(27%) | 29/53<br>(53%) | 26/34<br>(76%) | 9/10<br>(90%) | 7/8 (88%) | 0.003 |
| Other drugs<br>n (%) | 39/115<br>(34%) | 1/11<br>(9%) | 14/52<br>(27%) | 14/34<br>(41%) | 6/10<br>(60%) | 4/8 (50%) | NS |
| GH (ng/ml) | 14,5<br>(0.5-135) | 7,8<br>(1.3-40) | 12,8<br>(2.8-130) | 26<br>(0.5-135) | 33<br>(3.5-40) | 39<br>(12-100) | 0,036 |
| IGF-1 (ng/ml) | 734<br>(341-2197,0) | 634<br>(429-993) | 734<br>(439-1359) | 748<br>(341-2197) | 699<br>(439-1261) | 891<br>(608-1155) | NS |
| IGF-1 (x ULN) | 2.7<br>(1.2-6.1) | 2.5<br>(1.9-4.1) | 2.8<br>(1.4-5.3) | 2,6<br>(1.2-6.1) | 3<br>(1.7-4.1) | 2.9<br>(2-5.7) | NS |
| Surgical control<br>n (%) | 29/84<br>(35%) | 3/5<br>(60%) | 17/37<br>(46%) | 7/28<br>(25%) | 1/8<br>(12%) | 1/6 (17%) | NS |
| Disease control<br>n (%) | 70/105<br>(67%) | 6/8<br>(75%) | 33/49<br>(67%) | 18/31<br>(58%) | 8/9<br>(89%) | 5/8 (62%) | NS |

SRL, somatostatin receptor ligands; ULN, times of upper limit of normal

### Comparison among the five groups categorized according to the adenoma size at diagnosis

The prevalence of hypopituitarism, invasiveness and need for SRL differed between the five groups evaluated (p < 0.01), as did the initial GH levels (p = 0.036). There was no statistically significant difference in relation to male or female prevalence, presence or duration of symptoms, hypertension, and type 2 diabetes mellitus, T2 signal on MRI, need for RTX, other medications, IGF-1/ULN levels, cure surgery or current disease control. A positive correlation was found between adenoma size and hypopituitarism (r=0,517, p<0.01), adenoma size and invasiveness (r=0,457, p<0.01) and postsurgical control and disease control (r = 0.413; p <0.01).

### Comparison between adenomas < 10 mm vs ≥ 10 mm

Patients with microadenomas were older than the others (51 ± 10 yrs vs 42 ± 13 yrs; p = 0.02), had a lower prevalence of hypopituitarism (0% vs 31%; p = 0,03), invasive adenomas (0% vs 45%; p < 0,01), had lower GH levels (7.8 ng/ml vs 17 ng/ml; p=0,03) and needed less SRL (27% vs 67%; p = 0,02).

### Comparison between Groups II and III

Patients in Group II had a lower prevalence of hypopituitarism (9.8% vs 47%; p < 0.01), needed less SRL (53% vs 76%; p = 0.041) and radiotherapy (2% vs 15% ; p = 0.03). Patients in Group III had a lower proportion of hypointense adenomas and a higher proportion of T2 hyperintense adenomas (30% vs 63% and 26% vs 7%; p= 0.047).

### Comparison between Groups II and IV

Patients in Group II had a lower prevalence of hypopituitarism compared to Group IV (9.8% vs 50%; p < 0.01) and needed less SRL (53% vs 90%; p = 0.037). Patients in group IV had a lower proportion of hypointense adenomas (0% vs 63%; p = 0.011), and a higher proportion of isointense and hyperintense adenomas on T2-weighted MRI (83% vs 30% and 17% vs 7%; p = 0.011).

### Comparison between Groups II and V

Patients in Group II had a lower prevalence of hypopituitarism (9.8% vs 86%; p < 0.01), invasive tumors (33% vs 86%; p = 0.011) and needed less radiotherapy (2% vs 25 %; p= 0.041). Furthermore, patients in Group V had higher GH levels compared to Group II (38.6 ng/ml vs 7.8 ng/ml; p = 0.012).

### Comparison between Groups III and IV

There were no statistically significant differences in the comparison between the groups, whether in quantitative or qualitative variables.

### Comparison between Groups III and V

There were no statistically significant differences in the comparison between the groups, whether in quantitative or qualitative variables.

### Comparison between Groups IV and V

There were no statistically significant differences in the comparison between the groups, whether in quantitative or qualitative variables.

### ROC Curve

ROC Curve analysis was performed with the aim of evaluating the sensitivity and specificity of tumor size in predicting the various clinical outcomes that showed significance in comparisons between study groups. To evaluate hypopituitarism, data from 111 patients were analyzed, AUC = 0.818 [CI (95%): 0.734-0.902, p < 0.01], with a cutoff point of 19.5 mm of tumor diameter, presenting a sensitivity of 83. 9% and specificity of 70.5% in predicting the presence of hypopituitarism. Invasiveness was evaluated in 111 cases, AUC = 0.768 [CI (95%): 0.680-0.855, p < 0.01], with tumors greater than or equal to 17.5 mm demonstrating sensitivity of 73.3% and specificity of 61.5% to predict this outcome. The analysis of radiotherapy treatment included 117 patients, AUC = 0.784 [CI (95%): 0.673-0.895, p < 0.01], with a tumor diameter cutoff point of 20.5 mm demonstrating sensitivity of 88.9 % and specificity of 71.7% for the need for radiotherapy. Drug treatment with SRL was analyzed in 117 patients, AUC = 0.724 [CI (95%): 0.628-0.822, p < 0.01], sensitivity of 71.2% and specificity of 65.9% for tumors equal to or greater than 16 mm. The use of other medications was analyzed in 115 patients with an AUC of 0.644 [CI (95%): 0.537-0.752, p = 0.012], tumors equal to or greater than 19.5 mm showing a sensitivity of 61.5% and specificity of 62.2% in predicting the use of other medications. Analysis of the surgical control showed an AUC = 0.650 [CI (95%): 0.529-0.775, p = 0.002], with a tumor diameter of 20.5 mm or more demonstrating sensitivity of 81.5% and specificity of 47.3 % in predicting worse prognosis after surgery (**Figure 1**).

**Figure 1.**
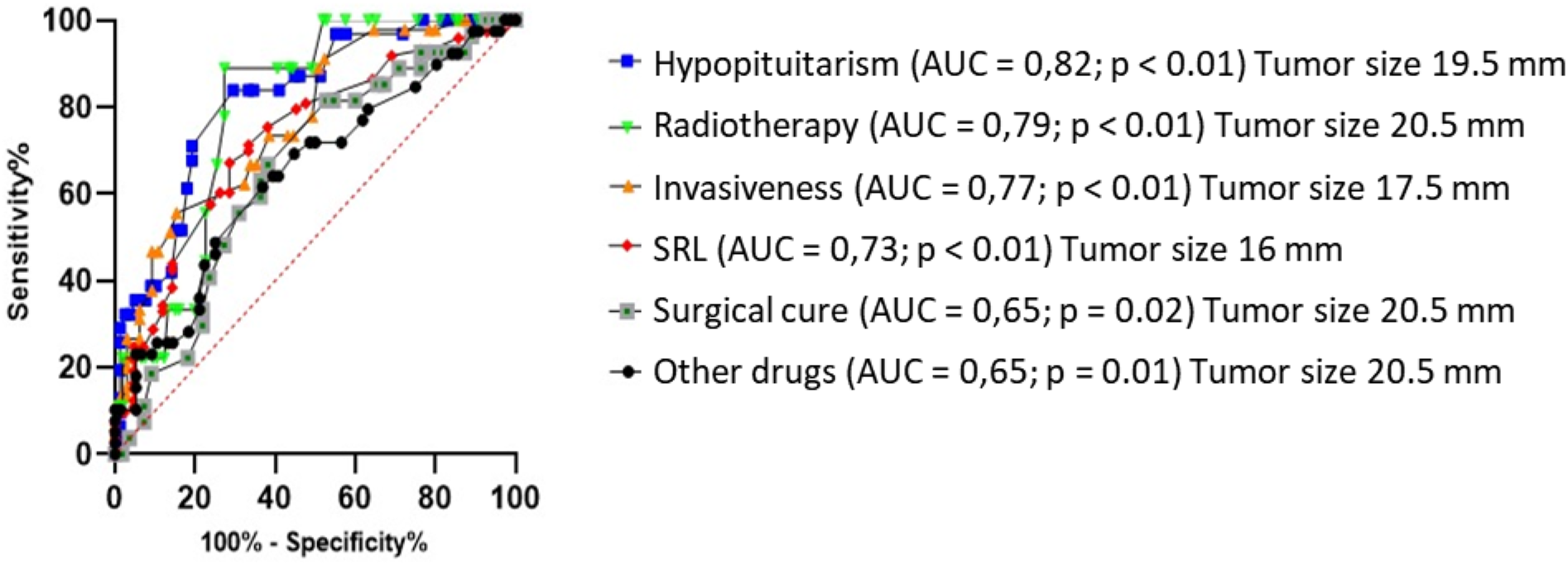
ROC Curve to evaluate tumor size in predicting the various clinical outcomes studied.

## DISCUSSION

Pituitary adenomas are traditionally classified according to their size at diagnosis in micro or macroadenomas using a cutoff of 10 mm (1 cm), which has been associated with differences in clinical presentation, management, and prognosis. Giant pituitary adenomas are usually defined as those equal to or greater than 4 cm in one plane, and they tend to be more invasive and associated with more morbidities, worse surgical and medical therapeutic responses and poor prognosis [3 - 5, 8]. However, there are scarce evidence that clinical signs, prevalence of morbidities, response to surgery and medical therapies and prognosis of giant adenomas as currently defined differ from smaller macroadenomas.

In our study we observed that, although some outcomes such as hypopituitarism, invasiveness, and the need to use SRL present a positive correlation with the increase in tumor size, other outcomes were better discriminated when the cutoff point used was 20 mm (as need treatment with radiotherapy or other medications in addition to SRL, hyperintense signal on T2, and cure with surgery). Furthermore, when we compared the groups of macroadenomas larger than 20 mm with each other (groups III, IV and V of the study), we did not observe statistically significant differences in any of the variables, whether qualitative or quantitative. This suggests that from a tumor diameter at diagnosis equal to or greater than 20 mm, the clinical, biological behavior and therapeutic and prognostic responses do not differ substantially. Analysis of the ROC curves demonstrated that most outcomes were related to a cutoff point of around 20 mm, indicating that microadenomas and macroadenomas smaller than 20 mm present a more favorable behavior than larger adenomas; but from 20 mm onwards the behavior is similar, with no difference between macroadenomas between 20-39 mm compared to giant adenomas.

In our series we found a prevalence of 7% of adenomas equal to or greater than 40mm, a higher prevalence than that reported in the literature by Shimon et al. [4], of less than 5%, but in accordance with what was reported by other studies, between 5 and 10% [9]. Our, patients with giant adenomas were slightly older than in the largest published series of patients with giant GH-secreting pituitary adenomas [4]. However, as observed in other cohorts, the group harboring microadenomas were significantly older compared to patients with macroadenomas [2910, 11].

The prevalence of hypertension and type 2 diabetes mellitus, which are comorbidities classically associated with acromegaly, were 49% and 25%, respectively. The prevalences found are within those reported in the literature by other authors [12 - 15].

Hypopituitarism was present in 28% of our patients, and the prevalence clearly increased with the size of the adenoma, going from 0% in Group I to 10% in Group II and reaching 47%, 50% and 86% in patients in Groups III, IV and V, respectively. Similarly, invasiveness, which is considered a very relevant prognostic factor in relation to achieving a disease-free state and the risk of recurrence or progression after surgery, was also associated with adenoma size [16 - 21]. Of note, both prevalence of hypopituitarism and invasiveness did not differ in the group of macroadenomas, with tumors ≥ 20 mm presenting similar frequencies compared to the larger ones.

The histological pattern of GH-secreting adenomas based on the cytoplasmic distribution of immunoreactive cytokeratin classifies these lesions in densely or sparsely granulated, [1012]. Densely granulated adenomas generally correspond to smaller lesions, are more frequent in older patients and respond better to SRL, while sparsely granulated adenomas correspond to larger, more aggressive tumors, more frequently occurring in younger patients and are associated with a worse response to SRL and prognosis [22 - 24]. In addition, T2-weighted MRI signal intensity may predict the histological characteristics of GH-secreting adenomas; as T2-hypointense lesions generally correspond to densely granulated adenomas and T2-hyperintense lesions generally correspond to sparsely granulated adenomas [23 - 25]. Unfortunately, we were unable to evaluate the histological characteristics of our patients’ adenomas, as this data was unavailable in most of the cases. However, based on the radiological examination of the pituitary, our data agree with the radiological-histological relationship, and we observed a higher prevalence of hypointense adenomas in Groups I and II and a higher prevalence of hyperintense lesions in Groups III, IV and V, but with no difference when the comparison was between the macroadenomas of these groups. The same was observed in relation to the need for radiotherapy, with patients with tumors ≥ 20 mm requiring radiotherapy in a greater proportion than patients with smaller tumors, but again with no difference in the adenomas ≥ 30 mm or ≥ 40 mm. In relation to drug therapy with SRL, its frequency increased with the size of the tumor, however, among patients in Groups III, IV and V, no difference was observed. The need for complementary drug therapies to SRL (cabergoline, pegvisomant, cabergoline and pegvisomant or bromocriptine) was also greater in patients with tumors equal to or larger than 20 mm.

Tumor size is one important factor in determining surgical outcome, regardless of invasiveness, and initial GH levels appear to correlate negatively with the surgery outcome [3, 27, 27]. This was confirmed in our series, as patients with adenomas smaller than 20 mm had a higher prevalence of disease control after surgery than patients with larger adenomas. Furthermore, we also found a positive correlation between post-surgical control and disease control. Of note, there was no difference in rates of disease control after surgery among adenomas larger than 20 mm, which is in agreement with the study by Koylu et al. [2828], where similar remission rates were observed when the cutoff of 20 mm was employed. In a series of 98 patients, Shimon et al. [2727] found a remission rate after surgery of 73% in tumors with 11-20 mm, but only in 20% of patients with tumors equal to or larger than 21 mm. Similarly, Ahmed et al. [2626] also found a significant difference in remission rates after surgery comparing patients with microadenomas (remission rate: 91%) mesoadenomas (tumor between 10-20 mm; 80%) and macroadenomas (tumors ≥ 20mm; 45%).

The main limitations of the present study include its retrospective design and the small number of patients in the categories of microadenomas and giant adenomas. Despite these limitations, we believe that our results open a venue to better characterization of giant GH-secreting adenomas, not only based on their size at diagnosis, but also in relation to their behaviour.

Therefore, our study suggests that a macroadenoma smaller than 40 mm and larger than 20 mm may have the same clinical, therapeutic, and prognostic implications as macroadenomas classically defined as giant, based solely on their size. If these results are confirmed in larger multicenter studies, we believe that the definition of giant adenomas should be revised to consider not only the size of the adenoma at the time of diagnosis, but also considering its clinical and biological behavior, therapeutic and prognostic outcomes.

## Data Availability

All data produced in the present study are available upon reasonable request to the authors

## Compliance with Ethical Standards

-Disclosure of potential conflicts of interest: the authors have nothing to declare.

-The authors received no financial support for the research, authorship, and/or publication of this article.

